# Partial Anchored Capture and Long-Read Sequencing (PACLseq) Enable a Stand-alone Diagnostic Method for Ph-like Acute Lymphoblastic Leukemia

**DOI:** 10.1101/2023.08.19.23294152

**Authors:** Hang Zhang, Huan Yu, Yanmei Chen, Kai Jiang, Beibei Huo, Jialin Li, Ting Liu, Dan Xie

## Abstract

Fusion genes play a crucial role in the development of Philadelphia chromosome–like acute lymphoblastic leukemia (Ph-like ALL). Timely and accurate determination of malgenic fusion transcripts that cause Ph-like ALL is essential for guiding treatment decisions. However, due to the complexity of possible gene fusion combination of Ph-like ALL, prevailing molecular diagnostic methods for Ph-like ALL are inefficient and lack of standardization, resulting in a slow diagnostic process. We introduce Partial Anchored Capture and Long-Read Sequencing (PACLseq), a nanopore-sequencing-technology-based approach, which enables fast stand-alone identification of fusion genes with a mere 10ng of input RNA. With extensive testing using BCR-ABL1 standards and 47 clinical samples to validate the efficacy of PACLseq, we demonstrated that PACLseq performs excellently in target region coverage and fusion gene detection accuracy, achieving a sensitivity of 93.33% and specificity of 100%. These findings highlight the reliability and versatility of PACLseq as a streamlined method for the clinical diagnosis of Ph-like ALL. By offering rapid and accurate fusion gene detection, PACLseq has the potential to significantly improve diagnostic efficiency, facilitate timely treatment decisions, and enhance patient outcomes in the management of Ph-like ALL.

## Introduction

Philadelphia chromosome-like acute lymphoblastic leukemia (Ph-like ALL) was initially described in 2009 by Mullighan’s group in the United States [1] and Den Boer’s group in the Netherlands [2]. These two research groups independently identified a novel subtype of ALL that lacked the Philadelphia chromosome but exhibited gene expression profiles similar to Philadelphia chromosome-positive ALL (Ph+ ALL). This subtype also showed partial sensitivity to tyrosine kinase inhibitors (TKIs) and had a poor prognosis. Consequently, this newly identified subtype was designated as Ph-like ALL. In 2016, the World Health Organization (WHO) officially classified Ph-like ALL as a new high-risk subtype of B-cell acute lymphoblastic leukemia (B-ALL) [3].

Ph-like ALL is characterized by highly complex genetic alterations, primarily involving fusion genes. These fusions can be classified into following subsets: ABL family (ABL1, ABL2, CSF1R, and PDGFRB), JAK/STAT pathway-related genes (CRLF2, JAK2, and EPOR), and other kinases (BLNK, DGKH, FGFR1, IL2RB, LYN, NTRK3, PDGFRA, PTK2B, and TYK2) [4]. Each of these genes may have different breakpoints and partner genes, leading to distinct fusion transcripts. Currently, a total of 83 fusions have been reported [5–20], and it is anticipated that more fusions will be discovered in the future with the advancement of next-generation sequencing (NGS) technologies. The detection of fusions serves not only as a diagnostic tool but also as a foundation for subsequent targeted therapy decisions. In Ph-like ALL patients with ABL family fusions, similar to Ph+ ALL, TKIs have demonstrated efficacy. Previous research has shown that imatinib and dasatinib can induce remission and eliminate minimal residual disease in patients with ABL-class fusion who exhibit an inadequate response to chemotherapy [5, 21–24]. Furthermore, studies have indicated that the addition of ruxolitinib to post-induction chemotherapy can lead to complete or partial remission in chemotherapy-resistant or relapsed Ph-like ALL patients with JAK2 or EPOR fusions[5, 25–29].

In summary, achieving an accurate diagnosis and implementing targeted therapy is pivotal for the effective management of Ph-like ALL at the time of initial diagnosis. Currently, three diagnostic protocols are employed for Ph-like ALL [30]: 1) The combination of flow cytometry and fluorescence in situ hybridization (FISH) method allows for the cost-effective and straightforward detection of a specific diagnostic panel. However, it has limitations in identifying fusion partners, not to mention fusions beyond the predefined panel. 2) Gene expression coefficients are calculated using a quantitative Real-Time Reverse Transcription Polymerase Chain Reaction (RT-PCR)-based low-density array (LDA) platform [31]. Patients who pass the screening undergo a series of downstream pathway gene analyses. This hierarchical diagnostic process is cumbersome and not well-suited for single-institution diagnosis. 3) The utilization of NGS techniques, such as whole exome sequencing, whole genome sequencing, or whole transcriptome sequencing (RNA-seq) [32], enables the comprehensive acquisition of molecular genetic information in patients. Nonetheless, due to the high cost, extensive data storage requirements, and intricate bioinformatics analysis involved, NGS has not been widely adopted in routine clinical practice. The aforementioned three methods typically have a diagnostic turnaround time of weeks, which falls short in meeting the imperative need for early diagnosis and timely targeted treatment of Ph-like ALL. Hence, the development of a rapid and accurate diagnostic approach is of paramount importance for Ph-like ALL management.

Targeted RNA sequencing represents a promising and cost-effective strategy for clinical applications. Notably, two studies have proposed that targeting either one partner or a partial short sequence of a transcript has the potential to capture the entire transcript, thereby enhancing the detection of new fusion transcripts. However, both of these concepts lack thorough validation[33, 34].

In an effort to streamline the diagnostic approach for Ph-like acute lymphoblastic leukemia (ALL) and capitalize on the advantages of long-read sequencing to identify fusions, including novel fusion partners, we have recently introduced an innovative methodology termed Partial Anchored Capture and Long-Read Sequencing (PACLseq). Built upon nanopore sequencing technology, PACLseq aims to identify fusion genes. This method facilitates the targeted analysis of a specific gene panel while simultaneously providing the opportunity to identify all fusion partner genes. In this study, we comprehensively evaluated the diagnostic prowess of PACLseq for the detection of fusions associated with Ph-like ALL.

## Result

### PACLseq method establishment and test

The PACLseq method is based on capturing a partial region of a complete or long fragment. In this study, our focus was on identifying fusion transcripts associated with Ph-like leukemia, specifically ABL2, CSF1R, PDGFRB, JAK2, ABL1, EPOR, and CRLF2 as target genes. We obtained transcript information for these genes from National Center for Biotechnology Information (NCBI) Refseq and merged the exon regions of all transcripts to create a unified region for target capture (Figure 1a and Supplementary Table 1). Biotinylated 100-mer probes were then designed with a ∼3.6X tiling density against the merged region by iGeneTech.

**Figure 1.**
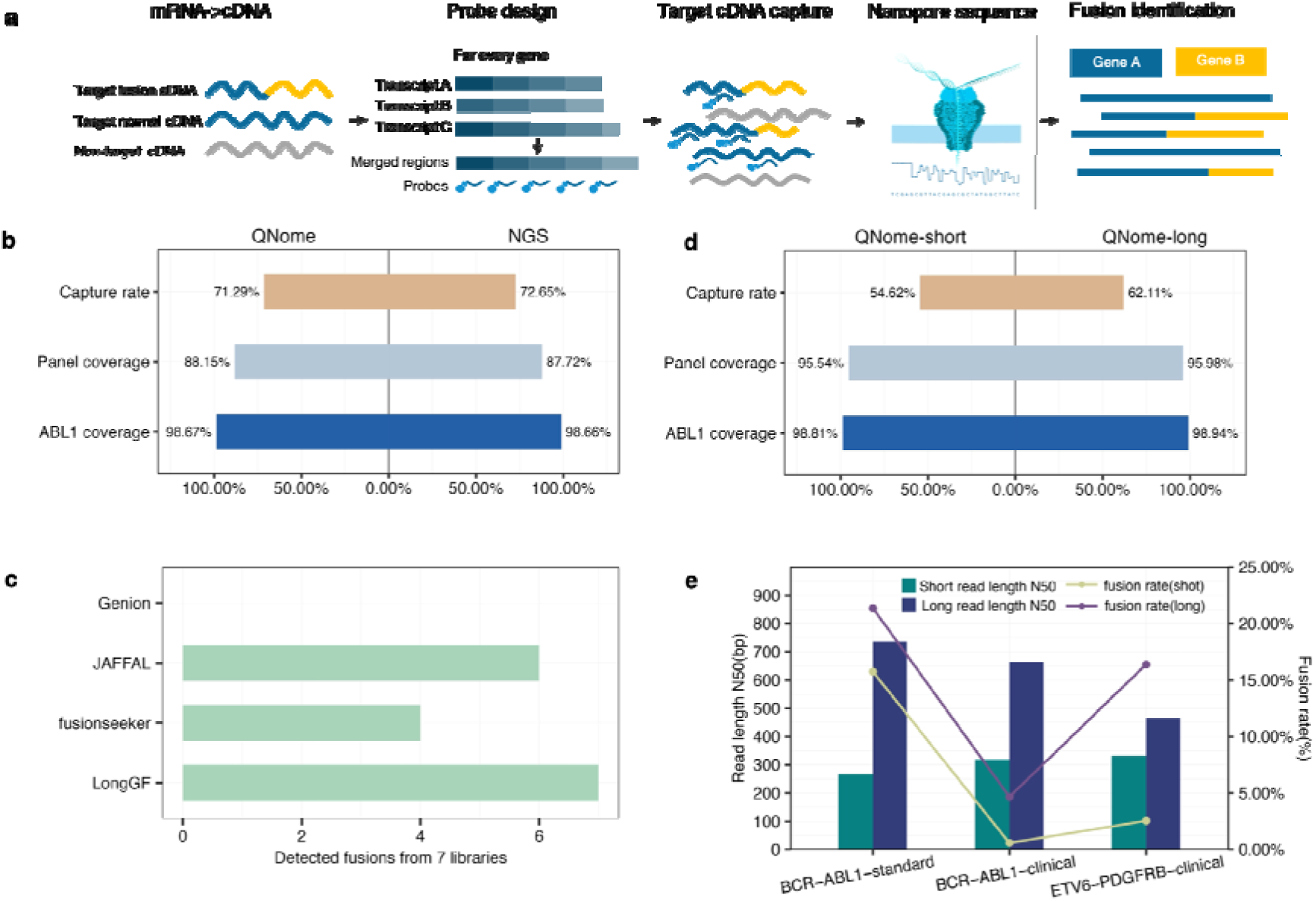
PACLseq scheme and performance. **a** PACLseq method scheme. The PACLseq method utilizes biotin-labeled probes designed for all transcripts, allowing for the capture of cDNA transcripts, including fusion transcripts. The captured cDNA transcripts are then subjected to nanopore sequencing. **b** Performance of BCR-ABL1 Standard capture and sequencing. The performance of BCR-ABL1 standard capture and sequencing was assessed on both the Illumina platform (left) and the QNome nanopore platform (right). **c** Capture performance of short-fragment and long-fragment nanopore sequencing. The capture performance of short-fragment sequencing (left) and long-fragment sequencing (right) on the nanopore platform was examined.**d** Evaluation of fusion detection software. Fusion detection software was evaluated, and LongGF successfully detected all 7 fusions. **e** Comparison of read-length and detected fusion rate. A comparison was made between the read length and detected fusion rate of short-fragment and long-fragment nanopore sequencing. The results suggested that longer read lengths may contribute to a higher fusion rate. This observation is supported by the analysis of the BCR-ABL1 fusion standard (left), clinical BCR-ABL1 fusion-positive sample (middle), and clinical ETV6-PDGFRB fusion-positive sample (right).

To assess the effectiveness of the PACLseq method, 2 short-fragment hybridization capture libraries (270-350bp) was constructed using the commercially available BCR-ABL1 standard sample. The library was subsequently sequenced using both Illumina and QNome nanopore platforms. The sequencing results showed excellent and comparable performance on both platforms. The capture efficiency exceeded 70%, with approximately 88% coverage and an average depth of over 2000x in the target region. Notably, ABL1 achieved coverage greater than 98% with an average depth exceeding 10,000x. Both platforms successfully detected BCR-ABL1 fusion, highlighting the effective capture performance of the panel ((Figure 1b, Supplementary Table 2, Supplementary Table 3).

### Validation of PACLseq and bioinformatics method establishment

To validate the PACLseq method and establish an effective bioinformatics analysis approach, we conducted experiments using clinically confirmed positive validation sample (1 BCR-ABL1 fusion and 1 ETV6-PDGFRB fusion). Short-fragment libraries (320-420bp) and long-fragment libraries (selecting fragments ≥200bp) were constructed from these samples and sequenced on the QNome platform. We also sequenced a long-fragment library of the commercially available BCR-ABL1 standard to compare with 2 previous results obtained from short-fragment sequencing.

To determine the most suitable fusion detection method, we evaluated several candidate software options, including LongGF [35], JAFFAL [36], Genion [37], and Fusionseeker [38]. After testing these methods on 7 libraries, we found that LongGF had the highest recall rate, successfully identifying fusion events in all 7 libraries. All those four methods contain some false positive fusions (Figure 1c, Supplementary Table 4). To optimize the analysis process and reduce false positive results, we developed a fusion analysis pipeline based on LongGF, which effectively detects fusions while minimizing false positives.

The results of our study demonstrated favorable coverage performance for both short-fragment and long-fragment sequencing of the 3 samples mentioned above. Capture rates ranged from 47.23% to 70.38%, with coverage of the target region ranging from 90.83% to 96.1% (Figure 1d, Supplementary Table 2). Moreover, among the 3 samples we detected, we observed that long-fragment sequencing outperformed short-fragment sequencing in fusion detection, showing a 5.28-fold higher fusion rate (Figure 1e, Supplementary Table 3).

These findings provide validation for the PACLseq method and establish a robust bioinformatics analysis approach for fusion detection. The use of long-fragment sequencing improves the accuracy of fusion detection and holds potential benefits for clinical applications. Our fusion analysis pipeline effectively identifies fusions while minimizing false positive results, enhancing the reliability of fusion detection in clinical samples.

### Evaluation of minimal cDNA input

In the context of limited RNA/cDNA availability in clinical samples, it is crucial to determine the minimal required cDNA input for PACLseq. The objective of this part was to identify the optimal cDNA input, as well as pre-capture and post-capture PCR conditions, for this sequencing method. To achieve this, a clinically confirmed positive validation sample (CCPVS-SFQ) was utilized, and different cDNA input levels were compared, including 100ng as the control and 50ng, 30ng, 20ng, and 10ng as the test groups. Long-fragment libraries (>200bp) were constructed using experimental combinations of pre-capture and post-capture PCR cycles, followed by sequencing on the QNome platform.

The performance of target region capture rate and coverage fell within the experiential range. Capture rates ranged from 39.60% to 67.49%, and coverage ranged from 95.86% to 95.94%. Notably, the ABL1 coverage at 500x ranged from 94.54% to 97.07%, and the average normalized depth of ABL1 varied from 1.02 to 1.17. Remarkably, all tested cDNA input levels successfully detected BCR-ABL1 fusion without any false positives, indicating the high effectiveness of using 10 ng cDNA input (Figure 2a, Supplementary Table 2, Supplementary Table 3).

**Figure 2.**
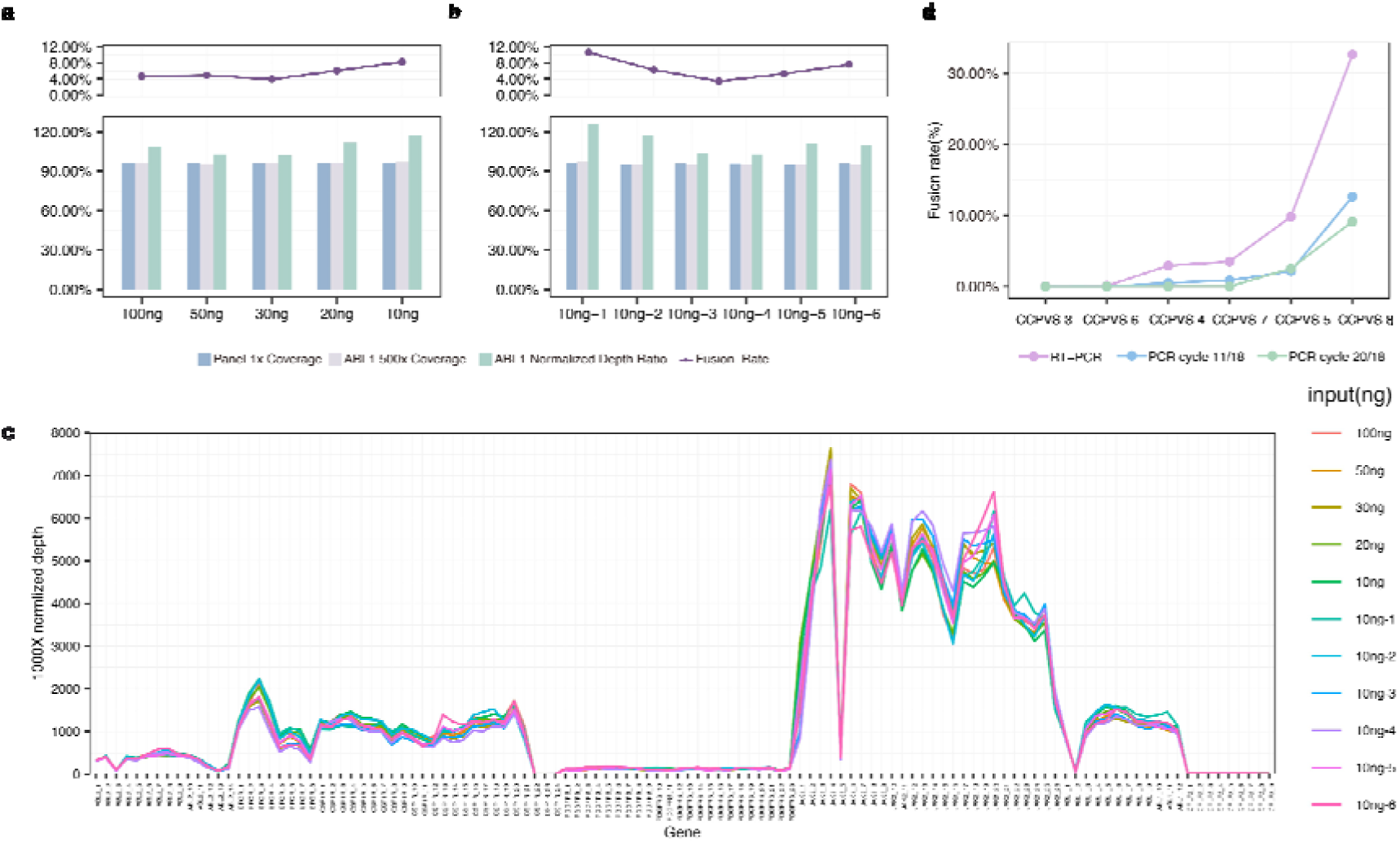
experiental parameter evaluate and clinical sample binding test. **a** Evaluation of minimal cDNA input. The minimal cDNA input was evaluated by testing 50ng, 30ng, 20ng, and 10ng inputs compared to the control (100ng) using a clinical BCR-ABL1 positive sample. The coverage of the target region, percentage of ABL1 coverage ≥500x, ABL1 average normalized depth ratio, and detected fusion rate were compared to the control. **b** Validation of performance with 10ng cDNA input.The performance of the method was validated using 10ng cDNA input with six repeats using different PCR cycles. All repeats showed comparable performance. c 1000x average normalized depth distribution of 11 input test libraries. **d** Optimal pre-captural and post-captural PCR cycle evaluation. The combination of 11/18 cycles showed highest fusion detection rate.

To further validate the performance of the 10ng cDNA input, additional 6 libraries were constructed using different PCR conditions and sequenced on the QNome platform.

These libraries consistently yielded BCR-ABL1 fusion results, with capture rates ranging from 30.79% to 51.5%, coverage ranging from 95.22% to 95.92%, ABL1 coverage at 500x ranging from 94.73% to 97.19%, and average normalized depth of ABL1 ranging from 1.03 to 1.26 (Figure 2b, Supplementary Table 2, Supplementary Table 3). Furthermore, the normalized depth at 1000x (1000*target gene average depth/panel average depth) for all eleven libraries exhibited a remarkably uniform distribution, implying the consistency of the 10ng input approach (Figure 2c).Among the tested combinations, the one with 11 pre-capture cycles and 18 post-capture cycles achieved the highest fusion rate at 10.63%, while other combinations yielded approximately 5% fusion rate.

Based on the consistent favorable coverage performance on ABL1 and the successful detection of the BCR-ABL1 fusion in all tests, we can preliminarily conclude that this method is suitable for testing with a 10ng input cDNA.

### Evaluation of optimal PCR conditions and detection limit of fusion rate

As we have determined that 10ng cDNA input works effectively in the PACLseq workflow, we proceeded to evaluate the optimal pre-capture and post-capture PCR cycles, as well as the fusion rate detection limit, using 6 clinically confirmed positive validation samples with fusion rates of 0.06%, 0.09%, 2.91%, 3.49%, 9.82%, and 32.65%. Based on the outcomes obtained from the preceding step, we identified two PCR conditions that exhibited favorable detection efficacy. Subsequently, we categorized the samples into two groups accordingly. One group underwent PCR cycles 11/18, which previously yielded the highest fusion rate of 10.63%, while the other group underwent PCR cycles 20/18, resulting in a fusion rate of 5.31% in the initial step.

The results demonstrated that the PCR cycle combination 11/18 yielded optimal results. Four out of six samples showed positive results, with the lowest RT-PCR detected fusion rate being 2.91% (0.72M reads) in the clinical setting. However, the other two samples with RT-PCR fusion rates of 0.06% (0.72M reads) and 0.09% (2.0M reads, ABL1 average depth is 4339.65x) were not detected. In contrast, the PCR cycle combination 20/18 only detected two out of the six fusions (Figure 2d, Supplementary Table 3). This further confirms the suitability of 10ng input cDNA in the PACLseq method, and we determined the pre-capture cycles to be 11 and the post-capture PCR cycles to be 18. Regarding the detection limit of PACLseq, we set the lowest fusion rate at 0.5%, calculated by dividing the number of fusion-supported reads by the average depth of the target captured gene breakpoint. We also set the minimum requirement of five breakpoint support reads in the bioinformatics pipeline. To meet these criteria, the sequencing depth must exceed 1000x. During this test, we successfully detected fusions with calculated fusion rates of 0.51% (with an RT-PCR fusion rate of 2.91%, supported by 14 reads out of 0.72M reads) and 0.85% (with an RT-PCR fusion rate of 3.49%, supported by 16 reads out of 0.86M reads). Based on our results, we hypothesize that our method can qualitatively detect fusion at a fusion rate as low as 10^-2^ level.

### Blind test validation using clinical samples

After configuring the key parameters in our method, including a fragment size of ≥200bp, a cDNA input of 10ng, and a PCR cycle combination of 11/18, we conducted a blind test using clinical blind-test samples. Since the current methodology of PACLseq can only identify coding regions, we omitted fusions involving intergenic regions in this part. In other words, we focused primarily on fusion genes other than IGH and IGK, and both partner genes should be confirmed in the clinical setting. Consequently, we categorized these samples into different groups based on the RIN values. Specimens with RIN > 3 were assigned to group A, while those with RIN ≤ 3 were designated as group B. Additionally, samples containing IGH, IGK-related fusions, or those with unknown partner genes were classified into group C.

The blind test included a total of 39 clinical samples, with 26 in group A, 9 in group B, and 4 in group C (Table 1, Figure 3a, Supplementary Table 5). These samples were sent to the laboratory for sequencing and analysis without revealing any information. The unblinding process was conducted after the acquisition of sequencing results.

**Figure 3.**
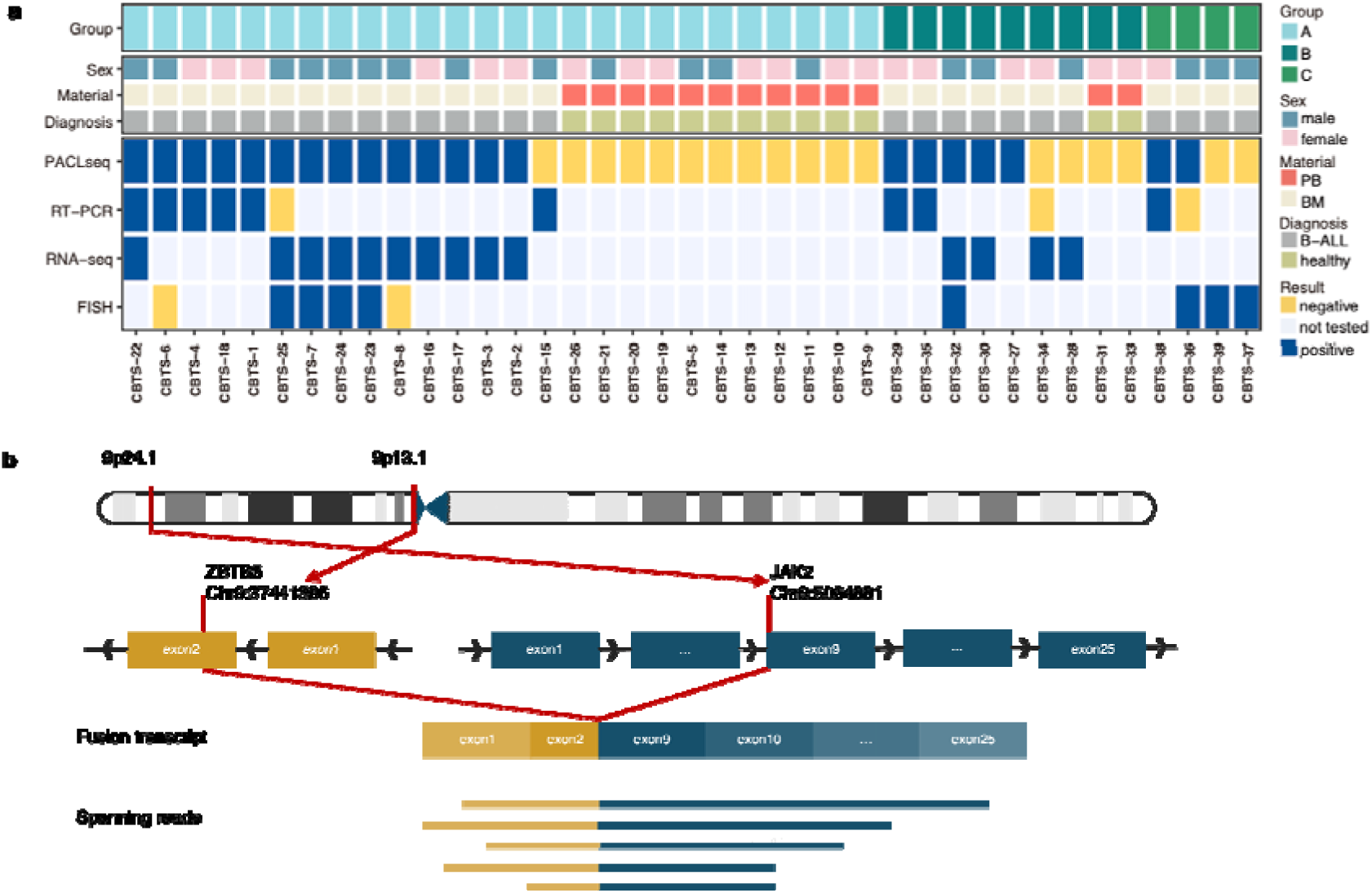
Fusion identification in clinical blind-test samples a Landscape of fusion gene identification in clinical blind-test samples, PB: peripheral blood, BM: bone marrow; b Structure of novel fusion gene ZBTB5-JAK2 identified in sample CBTS-25.

**Table 1.**
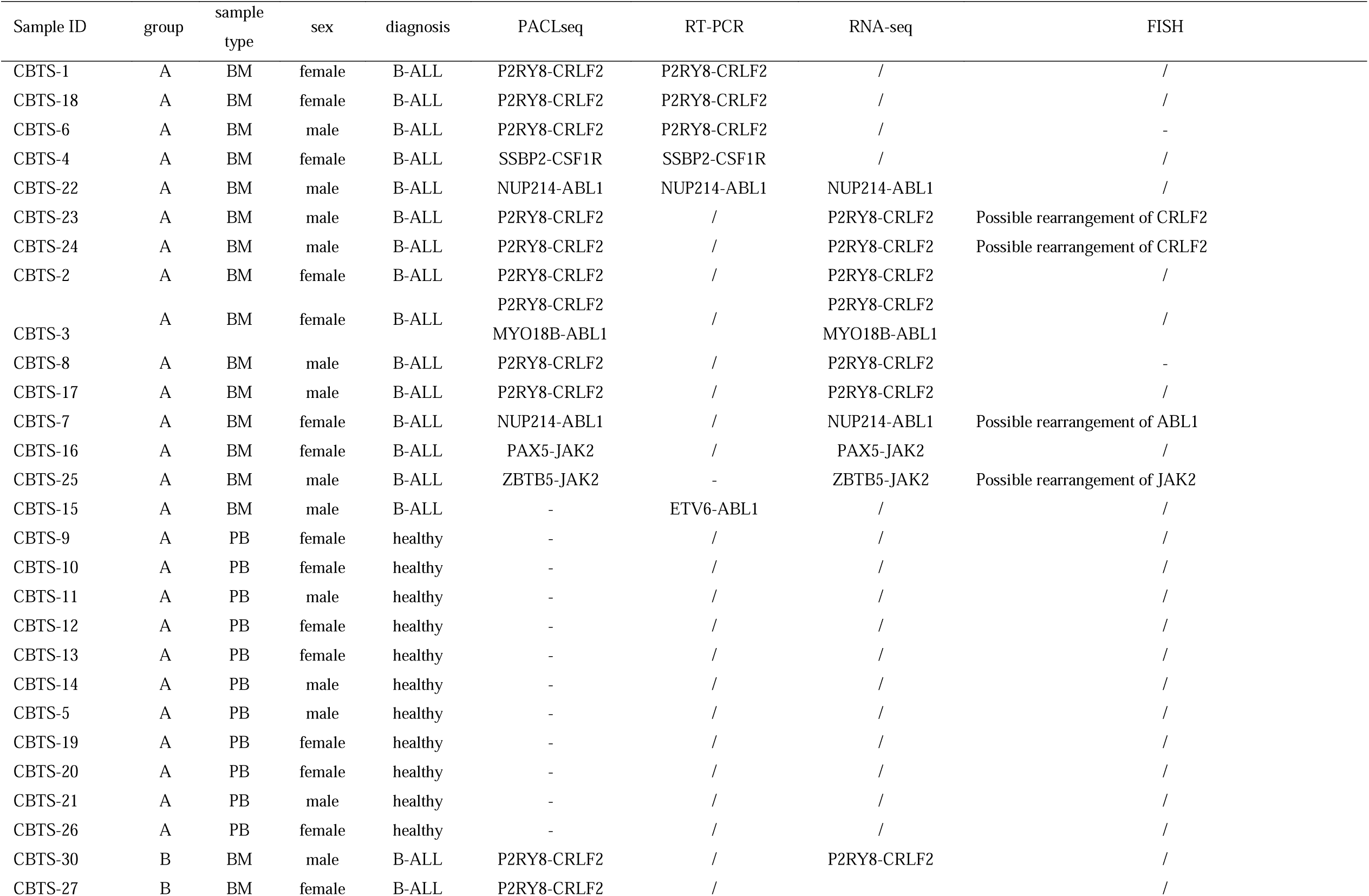
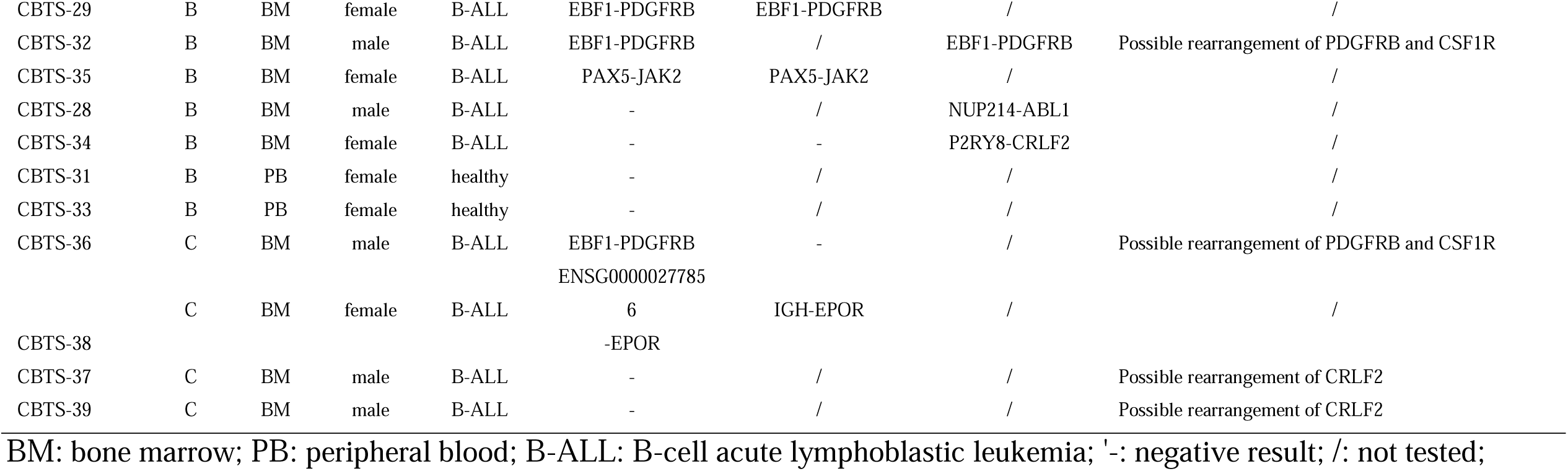
Basic information about clinical blind-test samples

Among the 26 samples in group A, 15 tested positive and 11 tested negative in the clinical setting. PACLseq achieved a recall rate of 93.33% and a precision rate of 100% for the positive samples, while the negative samples exhibited a specificity rate of 100%. In other words, one positive sample (CBTS-15) was not detected. Subsequently, we repeated the RT-PCR analysis on this sample, yielding a fusion rate of 0.3%. The PACLseq results for the remaining samples exhibited complete concordance with the clinical findings.

Among the 9 samples in group B, 7 tested positive and 2 tested negative. The positive samples in this group achieved a recall rate of 71.43% with a precision rate of 100%, and the negative samples exhibited a specificity rate of 100%. These results indicate that our method can still be effective for testing low-quality samples.

It is remarkable that we identified a new fusion, ZBTB5-JAK2 in sample CBTS-25 (Figure3 b), which further validates the ability of our method to detect novel fusion pairs. It is formed from exon 2 of ZBTB5 gene (ENST00000307750.5/NM_014872.3) (chr9: 37441386:-) and exon 9 of JAK2 gene (ENST00000381652.4/NM_004972.4) (chr9 5,064,881:+). After comprehensive search across various databases, including the TICdb database, the cancer genome atlas program (TCGA) fusion gene database, catalogue of somatic mutations in cancer (COSMIC) gene fusions database, ChimerDB 4.0 database[39] and Pubmed, we conclusively validated the novelty of the ZBTB5-JAK2 fusion.

In group C, PACLseq successfully detected an EBF1-PDGFRB fusion, where the partner gene information was not detected by RT-PCR. However, the other three samples were not detected, likely due to noncoding fusions which with significantly elevated target gene depth. In Group C, the clinical fusion detection was indeterminate, precluding the establishment of a gold standard. Consequently, we didn’t calculate diagnostic indicators. Furthermore, nanopore sequencing allows for the multiplexing of different samples on the same cell, reducing costs in clinical applications. To evaluate the multiplex performance on the QNome nanopore sequencing platform, we sequenced three samples that were indexed with different barcodes and pooled at equal molar concentrations. The analysis results remained consistent with the initial findings, confirming the reliability of multiplexing (Supplementary Table 5).

## Discussion

Our study devised a novel methodology, named PACLseq, based on nanopore sequencing technology. Subsequently, we developed a detection panel specifically designed for the identification of Ph-like ALL fusion genes. Through our research, we have successfully validated the diagnostic capability of PACLseq in detecting fusions within the designated panel, achieving this feat within a rapid turnaround time of 3 days. Notably, PACLseq exhibits exceptional performance in terms of recall, precision, and reproducibility, even when handling samples of poor quality. These outcomes underscore the diagnostic excellence of PACLseq in identifying Ph-like fusions. This holds immense significance within the diagnostic and therapeutic workflow of Ph-like ALL, addressing the critical need for early diagnosis and targeted drug intervention. Currently, numerous diagnostic strategies are employed in clinical applications to deal with complex fusion genes of Ph-like ALL (Table 2). With the exception of RNA-seq, most of these strategies often necessitate the combination of multiple detection methods. These approaches involve intricate steps, thereby increasing the complexity of clinical operations and placing additional burdens on patients. Regrettably, these approaches are confined to detecting fusions within a circumscribed range and lack the capacity to unveil novel fusions. While RNA-seq exhibits the potential to identify the complete spectrum of fusion genes. However, its operational intricacies render its widespread clinical adoption unfeasible. Due to the large scale of whole transcriptome, the sensitivity of RNA-seq in the detection of fusions is object to fusion gene expression levels and the commingling of normal tissue elements within the sample [40, 41]. In contrast, PACLseq has the capability to identify Ph-like fusions in a single test. This method offers high-quality results in a short turnaround time of 3 days. Furthermore, PACLseq targets Ph-like ALL gene panel (ABL2, CSF1R, PDGFRB, JAK2, ABL1, EPOR) at one end, thereby enhancing sequencing coverage for the focal gene, while simultaneously enabling the detection of the whole transcriptome at the other end. This innovative approach not only increases the sensitivity in the detection of fusions with the focal genes, but also facilitates the identification of novel fusion events. Our results demonstrated its capacity to discover novel fusions (CBTS-25), potentially expanding the panel of fusion genes associated with Ph-like ALL. Furthermore, the compact size and maneuverability of the QNome nanopore platform make it highly suitable as a single-institution solution. The modest throughput of PACLseq serves as an advantage in a single-institution platform, enabling immediate detection even for a single newly diagnosed patient during clinical practice. This eliminates the need to wait for adequate sample size or collaborate with other centers for large-scale testing. Consequently, PACLseq ensures timely results. Additionally, a single chip can accommodate the simultaneous testing of up to three samples, offering flexibility to address diverse scenarios involving newly admitted patients in clinical settings.

**Table 2.**
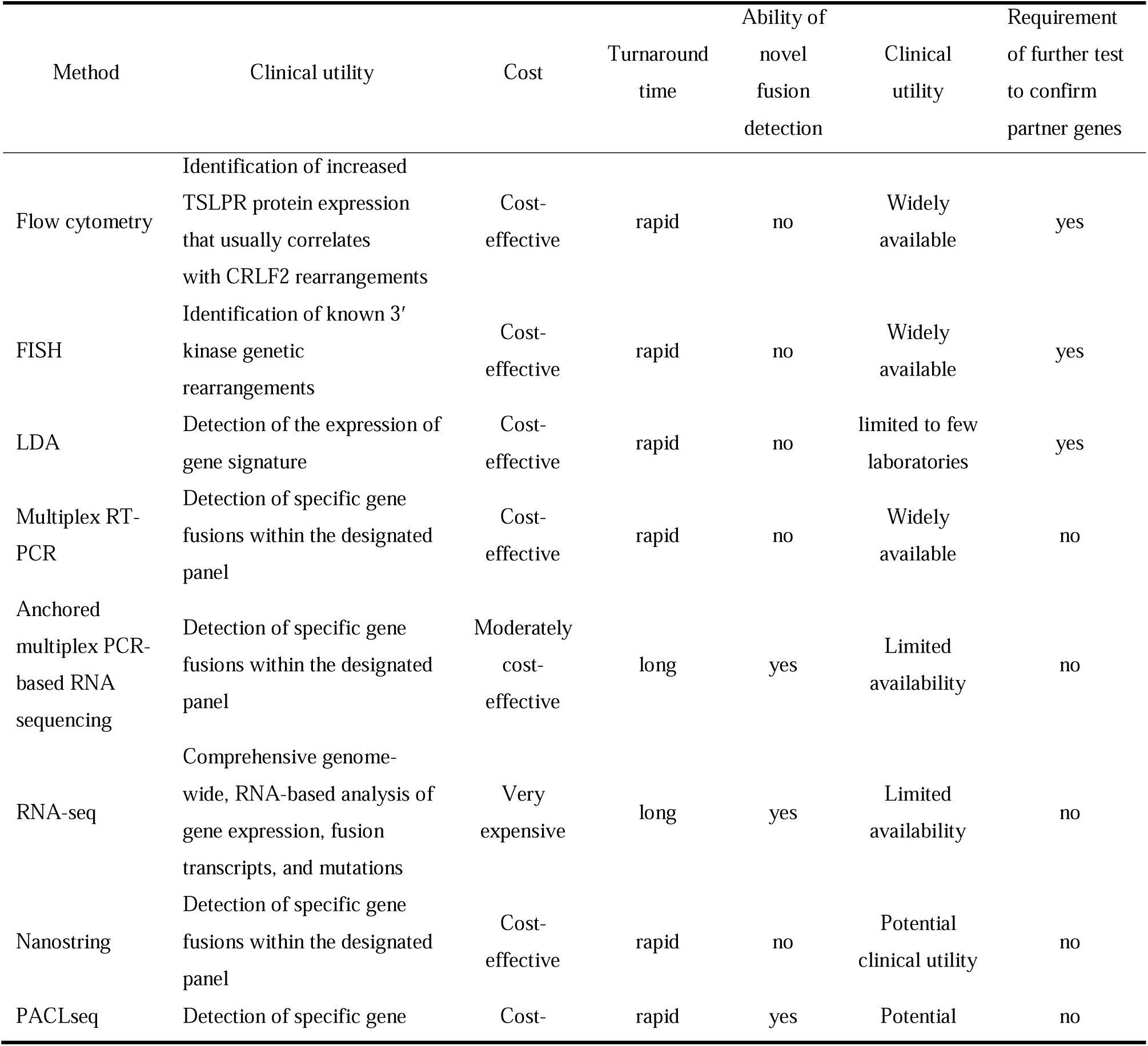
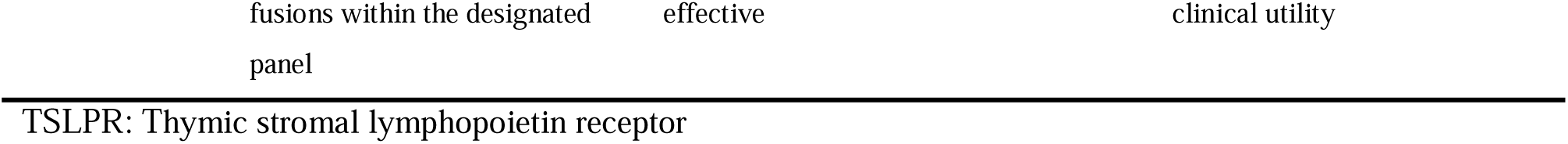
Summary of diagnostic tests used to determine the fusions related to Ph-like ALL

In this study, a series of repeated experiments were conducted on a clinically confirmed positive validation sample (CCPVS-SFQ) using varying cDNA input and PCR cycle conditions to investigate the optimal sequencing workflow. Interestingly, consistent positive results for the BCR-ABL1 fusion were obtained, and no false positive results were observed. This robust consistency confirms the reproducibility of the PACLseq. However, it is important to note that the fusion rate calculated by PACLseq did not correspond with the RT-PCR result observed in the clinical setting. Additionally, each repeated experiment exhibited distinct fusion rates, which can be attributed to two pivotal factors. Firstly, there is a time gap between the RT-PCR and PACLseq procedures, which may lead to RNA degradation and a decline in sample quality compared to fresh samples. Secondly, the fusion detection and calculation method employed in long-read sequencing is inherently less precise than the clinical PCR method. As a result, PACLseq is more suitable for qualitative analysis rather than quantitative analysis. Based on our comprehensive findings, we propose that PACLseq is capable of qualitatively detecting fusions at a fusion rate as low as 10^-2^ level, which is competent for diagnostic purposes.

RNA is highly unstable and susceptible to degradation. Presently, commercial NGS facilities commonly specify a prerequisite of an RIN ≥7 for subsequent steps of library construction and analysis. Previous investigations on degraded RNA have revealed the impact of sample degradation on gene expression. However, in samples with RIN >5, the influence of sample degradation on the proportion of affected genes is minimal, exerting an insignificant effect on the subsequent pathway enrichment results [42–45]. Nonetheless, the impact of RNA degradation on fusion gene detection is considerably more significant. In the context of fusion gene detection by RNA-seq, the efficacy to detect fusions is not only influenced by RNA degradation but also by the distance from the fusion breakpoint to the 3’ end of the gene. Even a slight decrease in RIN makes fusions challenging to detect when the distance exceeds 1 kb [46]. This effect is particularly notable in clinical specimens, where limited sample storage conditions prevail in clinical settings, leading to general RNA degradation. Furthermore, clinical single-institution platforms often lack the capability of performing RNA-seq and therefore rely on commercial NGS facilities. The transportation of specimens can contribute to a subsequent decrease in RNA quality, making fusion gene detection more challenging. However, our research demonstrates the remarkable tolerance of PACLseq towards sample quality. The method achieves a recall rate of 93.33% in samples with RIN >3. The only false negative observed could potentially be attributed to an extremely low fusion rate (only 0.3% detected upon retesting). Furthermore, PACLseq demonstrates a notable recall rate of 71.45% even in highly degraded specimens with RIN ≤ 3. Additionally, PACLseq requires a minimal input sample quantity of only 10 ng of RNA, thereby conserving sample material. This feature proves particularly beneficial for patients encountering challenges in obtaining bone marrow samples.

Although numerous computational methods exist for detecting fusions using short-read data, the availability of long-read gene fusion detection software is currently limited. In our study, we selected four software programs, namely LongGF [35], JAFFAL [36], Genion [37], and Fusionseeker [38], to validate fusion detection. A total of 7 tests were analyzed using these software programs. Notably, both LongGF and JAFFAL successfully detected most positive fusions, while Fusionseeker and Genion failed to detect true positive fusions in the majority of cases. It is important to highlight that LongGF and JAFFAL were validated using clinical leukemia samples, whereas the other software programs were solely tested using simulated data and cell lines. These findings emphasize the importance of incorporating clinical samples during the development phase of software to effectively validate its performance.

In our study, LongGF was ultimately chosen as the method for fusion detection. However, it is essential to acknowledge that LongGF may generate false positive fusions when dealing with homologous genes in the genome [35]. During our additional sequencing on sample “CBTS-38”, the clinical result using multiple RT-PCR indicated an IGH-EPOR fusion. However, the result obtained through LongGF was ENSG00000277856-EPOR, where ENSG00000277856 represents a homologous gene of IGHV3-43. This discrepancy can be attributed to the inherent limitations of the LongGF software. Another limitation of LongGF is its reliance on annotated transcripts. This means that fusions involving intergenic or intronic sequences at the breakpoint may go undetected.

Consequently, complex fusions such as IGH-CRLF2 might be missed. In our study, the sample “CBTS-37” and “CBTS-39” exhibited clinical FISH results suggestive of a CRLF2 rearrangement. Although PACLseq did not directly detect the corresponding fusion in this sample, it did reveal significantly elevated CRLF2 gene expression, which is commonly associated with CRLF2 gene rearrangements [47]. Therefore, even though PACLseq did not directly detect the fusion gene in this case, it indirectly suggests the presence of an IGH-CRLF2 fusion. Considering the limitations of LongGF in detecting fusions in non-coding regions, this indicates the potential presence of an IGH-CRLF2 fusion in the sample “CBTS-37” and “CBTS-39”. Unfortunately, due to limitations in both sample size and sample quality, we were unable to conduct additional experiments to verify our hypotheses.

In recent years, long-read sequencing has gained increasing recognition for their ability to enhance our comprehension of transcriptomic complexity compared to short-read RNA-seq. Short-read data, typically consisting of fragments less than 150bp in length, possesses inherent limitations in accurately detecting full-length gene isoforms. Short-read sequencing is susceptible when resolving repetitive or low-complexity regions.

Conversely, long-read sequencing generates reads spanning thousands of bases, facilitating the comprehensive capture of a majority of transcriptional isoforms within individual reads, and eliminating the need for transcriptome assembly. Our research demonstrates that when employing the PACLseq method for detecting fusion genes in clinically confirmed positive validation samples, long-read library exhibits a slightly higher capture rate and fusion rate compared to short-read library. Despite the relatively short fragment lengths generated by the current method (less than 1000bp) due to the poor quality of RNA, it has exhibited promising performance. Moving forward, optimization strategies can be explored to obtain longer fragments and conduct analyses in samples with better quality and larger-sample studies.

In conclusion, we have successfully established a novel detection method for identifying ph-like ALL-related fusion genes based on PACLseq. Through the validation of clinical samples, this method has demonstrated reliable detection results, along with a short turnaround time and resilience towards poor RNA quality and small sample input.

Consequently, it proves to be an excellent choice for diagnosis of ph-like ALL in clinical settings. In the future, we intend to conduct prospective studies using large sample cohorts to further evaluate the reliability of PACLseq in diagnosing ph-like ALL, as well as explore the performance of long-read sequencing for fusion gene detection. Additionally, we plan to design other panels to investigate further clinical applications of PACLseq.

## Method

### Standard and clinical samples

Commercially available BCR-ABL1 standard sample was purchased as RNA format (BCR-ABL1,P210 Fusion, CBP20061R, COBIOER BIOSCIENCES CO., LTD]. The collection of patient samples was ethically approved by the Biomedical Ethics Committee of West China Hospital (approval number: 2023-390). Clinically confirmed positive validation samples were obtained from the peripheral blood or bone marrow, and the fusion rate was quantified using RT-PCR in the clinical setting. The clinical blind-test negative samples were collected from the peripheral blood of healthy donors, while the positive samples were obtained from the bone marrow of first diagnosed B-ALL patients. The presence of ph-like fusion genes was clinically confirmed using FISH, multiplex RT-PCR, or RNA-seq methods performed in Kindstar Globalgene Technology, Inc. RNA extraction from both bone marrow and peripheral blood samples was performed using the TRIzol-based RNA extraction method.

### Commercial probe synthesis

The transcript information for these genes was initially obtained from NCBI Refseq. Subsequently, the exon regions of all transcripts were merged for each gene to establish a merged region for target capture(Supplementary Table 1). Commercially synthesized probes were designed as biotinylated 100-mers with a ∼3.6X tiling density against the merged region by iGeneTech (iGeneTech, T040V19). And the target capture kit TargetSeq also provided by iGeneTech (iGeneTech, C10831).

### Library construction

The size distribution of mRNA was assessed using the Agilent Tapestation 4150 system, and samples with a RIN (RNA Integrity Number) value of ≤ 3 were determined to have undergone significant degradation. The mRNA samples were then chemically fragmented, followed by the synthesis of double-stranded cDNA. Subsequently, the cDNA was utilized for library construction, which included pre-capture library construction, target enrichment, and post-capture library construction, according to the instructions provided by the manufacturer (mRNA-seq Lib Prep Kit for Illumina, RK20302, ABclonal). For target enrichment, the pre-capture library was subjected to capture using the iGeneTech TargetSeq One® Hyb & Wash Kit with Eco Universal Blocking Oligo (iGeneTech, C10732) following the provided protocol. For more detailed information, please refer to the supplementary methods.

### QNome-3841 nanopore sequencing

After the post-capture library construction, the library product was prepared for nanopore sequencing. Specifically, the product (300fmol) underwent end-repair and nanopore adapter ligation using the QNome-3841 library preparation protocol (QLK-V1.1.1, QitanTech). Subsequently, the library product (80fmol) was loaded onto the QNome-3841 nanopore cell for sequencing.For illumina short-fragment sequencing, the libraries were sent to Novogene for sequencing on the Novaseq6000 platform. For more detailed information, please refer to the supplementary methods.

### Bioinformatics analysis

We have developed a streamlined bioinformatics workflow for analyzing QNome nanopore data, which can be found at https://github.com/HuanYuu/TargetFusion. This workflow incorporates quality control, reference alignment, fusion detection, and statistical analysis. The sequenced fastq files underwent filtering using nanofilt to remove reads with lengths shorter than 100bp or quality scores lower than 7. The filtered reads were then aligned to the ensemble GRCh38 reference using minimap2 with the splice-aware option (-acx). Nanostat and mosdepth were used separately for subsequent calculations.

To identify fusions, we evaluated several candidate software tools, including LongGF, JAFFAL, Genion, and Fusionseeker. A BCR-ABL1 standard and four clinical positive samples, totaling nine libraries, were utilized for performance evaluation(Supplementary Method). Among the four software tools, LongGF exhibited the highest recall rate. The log files generated by LongGF, containing detailed read span information, were utilized for custom filtering to further improve precision. Consequently, LongGF was chosen for fusion detection, and the following filters were applied to the LongGF results:

1. The fusion gene pairs must be joined together in the 5’-3’ direction.
2. The average depth of the targeted gene’s breakpoint (20bp) must be greater than or equal to 100.
3. The fusion rate, calculated by dividing the number of fusion-supported reads by the average depth of the target captured gene breakpoint (20bp), must be larger than 0.5%.
4. The number of supported fusion reads must be at least 5.

The average 20bp breakpoint depth of the partner gene must be higher than the number of fusion-supported reads.

## Data Availability

The QNome nanopore sequence datasets data generated in this study have been deposited in the NCBI sequence read archive under accession number PRJNA1002848.

## Acknowledgements

We would like to thank Kindstar Globalgene Technology, Inc for their contribution in conducting fusion detection in the clinical setting and providing the results. We would like to thank all the participants in this study, including patients and nurses for their contributions. This work was supported by development fund from Department of Hematology, West China Hospital (ZX2021019).

## Author contributions

Ting Liu and Dan Xie conceived the project. Hang Zhang and Ting Liu provided patient samples and clinical data. Hang Zhang performed RNA extractions, Yanmei Chen and Beibei Huo performed library preparation and targeted sequencing. Huan Yu, Jialin Li and Kai Jiang performed bioinformatic analysis. Hang Zhang and Huan Yu wrote the manuscript with input from all authors.

## Data availability

The data supporting the findings from this study are included either in the manuscript or supplementary files. The QNome nanopore sequence datasets data generated in this study have been deposited in the NCBI sequence read archive under accession number PRJNA1002848. All data are also available from the authors upon reasonable request.

## Identifying information statement

The sampleIDs were not known to anyone outside the research group.

