## Supplementary Method for "Partial Anchored Capture and Long-Read Sequencing (PACLseq) Enable a Stand-alone Diagnostic Method for Ph-like Acute Lymphoblastic Leukemia"

#### PACLseq Protocol

PACLseq is a method designed to capture and sequence long-fragment cDNA. The process involves several steps, including:

1. RNA quantification and size distribution analysis;
2. RNA fragmentation;
3. cDNA synthesis;
4. pre-capture illumina library preparation;
5. target capture with a customized panel;
6. post-capture nanopore library preparation;
7. QNome nanopore sequence;
8. Bioinfomatic analysis;

**Mainly used reagent**:

Nuclease-free Water( AM9930, Thermo Fisher Scientific)
VAHTS® DNA Clean Beads(N411-03, Vazyme)
Equalbit 1×dsDNA HS Assay Kit(EQ121-02, Vazyme)
mRNA-seq Lib Prep Kit for illumina(RK20302, ABclonal)

- 2X Frag/Elute Buffer
- RT Reagent
- First Strand Synthesis Enzyme Mix
- Second Strand Synthesis Reaction Buffer
- Second Strand Synthesis Enzyme Mix
- Nuclease-free Water
- End-prep Buffer
- End-prep Enzyme Mix
- Ligation Buffer
- Ligase Mix
- 2X PCR Mix
- Low EDTA TE

RNA Adapter Module 96 Index for Illumina(RK20351/RK20352, ABclonal)
TargetSeq® Target Probes(iGeneTech, PT1008252)

TargetSeq® Target Probes
TargetSeq® Cap Beads & Nuclease-Free Water(iGeneTech, C10422)
TargetSeq® Cap Beads
Nuclease-Free Water
TargetSeq One® Hyb & Wash Kit with Eco Universal Blocking Oligo(iGeneTech, C10732)
TargetSeq One® Hyb & Wash Kit (Module A)
Hyb Human Block
RNase Block
TargetSeq One® Hyb Buffer
TargetSeq One® Hyb & Wash Kit (Module B)
Binding Buffer
TargetSeq One® Wash Buffer
Wash Buffer 1
TargetSeq One® Hyb & Wash Kit (Module C, for Illumina)
Post PCR Master Mix
Post PCR Primer (25 μM)*
TargetSeq® Eco Universal Blocking Oligo (for Illumina)
**QLK-V1.1.1(Qitan Tech)**
Sequencing adapter(SAC)
4x ligation buffer(LRB)
long fragment wash buffer(LWB)
short fragment wash buffer(SWB)
control DNA sequence(CDS)
elution buffer(AEB)
DNA ligaition enzyme(DLE)
DNA repair enzyme(DRM)
DNA repair buffer(DRB)
end repair enzyme(EPM)
**QSK-V1.1.1(Qitan Tech )**

### 1.RNA quantification and size distribution analysis

RNA quantification and size distribution analysis are conducted using the Agilent Tapestation 4150 system. Subsequently, 1ul mRNA is quantified using the qubit method.

### 2.mRNA fragmentation

The mRNA is chemically fragmented in 2X Frag/Elute Buffer at elevated temperatures. The fragmentation process is carried out under the following conditions:

| Component | 10ng/20ng/30ng/50ng/100ng |
|---|---|
| 2X Frag/Elute Buffer | 5ul |
| Total volume(add Nuclease-free Water) | 10ul |
| Processing | 94℃, 5min |

**3.cDNA synthesis**

After the mRNA fragmentation, the next steps involve first strand cDNA synthesis and second strand cDNA synthesis. These steps collectively enable the conversion of fragmented mRNA into double-stranded cDNA, facilitating further downstream applications such as library construction and sequencing.

First strand cDNA synthesis:

| Component | 10ng/20ng/30ng/50ng/100ng |
|---|---|
| mRNA(previous step) | 10ul |
| RT Reagent volume | 8ul |
| First Stand Synthesis Enzyme Mix | 2ul |
| Total volume | 20ul |
| Processing | 25℃, 10min;<br>42℃, 30min;<br>70℃, 15min; |

Subsequently, second strand cDNA synthesis is carried out, and the synthesized product is purified using VAHTS DNA Clean Beads to remove any unwanted impurities or residual reaction components.

| Component | 10ng/20ng/30ng/50ng/100ng |
|---|---|
| first strand cDNA product(previous step) | 20ul |
| Second Strand Synthesis Reaction Buffer | 8ul |
| Second Strand Synthesis Enzyme Mix | 4ul |
| Total volume(add Nuclease-free Water) | 80ul |
| Processing | 16℃, 2h |

| | |
|---|---|
| Purification(VAHTS DNA Clean Beads) | 1.8x |

**4.Pre-capture illumina library preparation**

After mRNA fragmentation, the resulting product undergoes library construction (Supplementary Figure 1). The cDNA product is subjected to end-repair and then ligated with an RNA Truncated adapter. Following this, a Universal PCR adapter is ligated to the product, and the amplified cDNA is purified. Subsequently, the target genes are captured and ligated with the nanopore adapter for further processing.

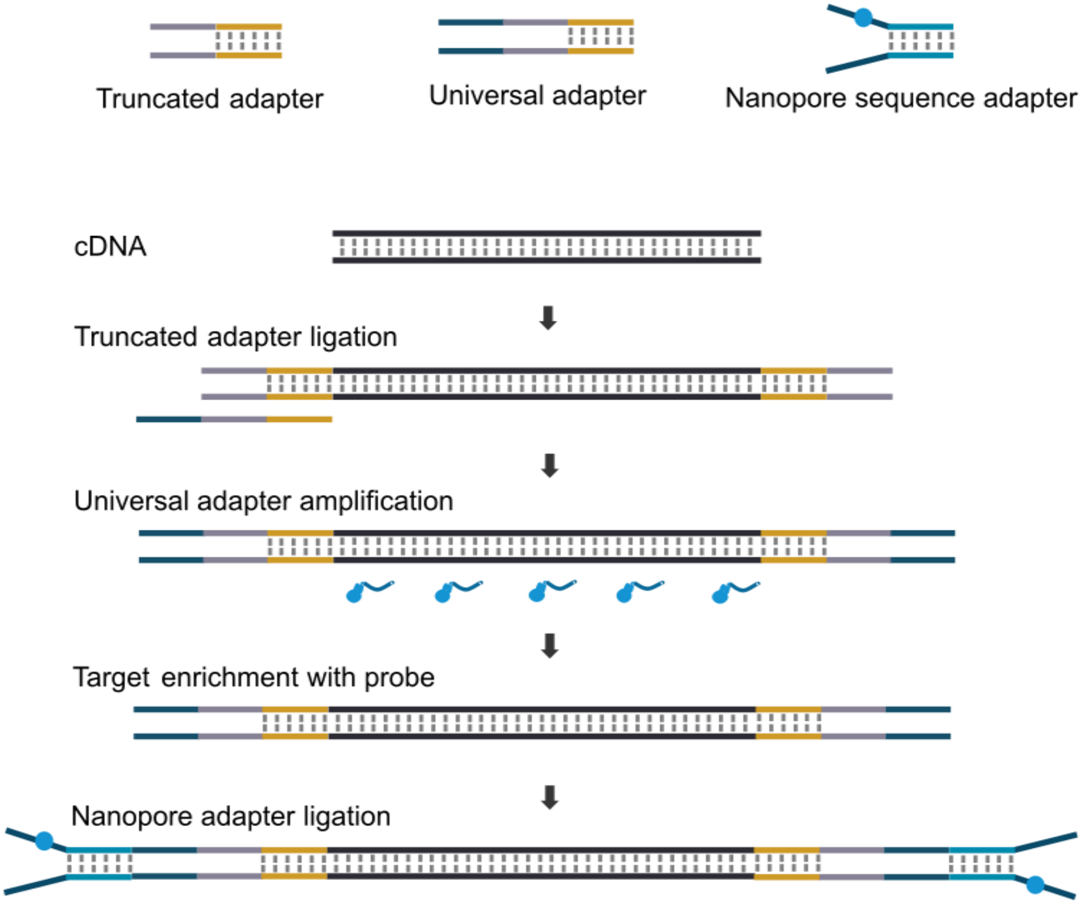


Supplementary Figure1: the scheme of library construction.

First, the cDNA fragments undergo end-repair, where any damaged or uneven ends are repaired to generate blunt ends. This is important for ensuring optimal ligation efficiency in the subsequent steps.

| Component | 10ng/20ng/30ng/50ng/100ng |
|---|---|
| Purified cDNA product(previous step) | 37ul |

| | |
|---|---|
| End-prep Buffer | 10ul |
| End-prep Enzyme Mix | 3ul |
| Total volume | 50ul |
| Processing | 30℃,30min;<br>65℃,30min; |

Next, the repaired cDNA fragments are ligated with an RNA Truncated adapter. The adapter contains sequences necessary for downstream processes.

| Component | 10ng/20ng/30ng/50ng/100ng |
|---|---|
| End-repaired an purified cDNA product(previous step) | 50ul |
| Ligation Buffer | 16.5ul |
| RNA Truncated Adapter | 2.5ul |
| Ligase Mix | 3ul |
| Processing | 22℃,30min; |
| Purification(VAHTS DNA Clean Beads) | 0.45x |

Following the ligation with the RNA Truncated adapter, a Universal PCR adapter is ligated to the cDNA fragments and purified. This adapter allows for the amplification of the cDNA fragments in the subsequent PCR step.The ligated product, containing both the RNA Truncated adapter and the Universal PCR adapter, is subjected to PCR amplification using primers specific to the universal adapter sequences. After PCR amplification, the amplified product is purified and ready for the subsequent target capture step.

| Component | 10ng/20ng/30ng/50ng/100ng |
|---|---|
| RNA Truncated adaptered product (previous step) | 20ul |
| 2×PCR Mix | 25ul |
| Universal PCR Primer | 2.5ul |

| | |
|---|---|
| PCR Index Primer | 2.5ul |
| PCR amplification processing | 98℃45s;<br>98℃ 10s;<br>60℃ 15s;<br>72℃ 1min; |
| PCR cycle(different cycle for different input) | 10ng-1:11<br>10ng-2:20<br>20ng:18<br>30ng:18<br>50ng:16<br>100ng:13 |
| Processing | 72℃ 5min; |
| Purification(VAHTS DNA Clean Beads) | 1x |

**5.Target capture with a customized panel**

Target capture with a customized panel is conducted by setting up a reaction system with the appropriate conditions. The previous product is quantified, and 750ng of the product is concentrated for subsequent use. The mixture is gently mixed and incubated for 3 minutes. Subsequently, 28ul of the supernatant is combined with 2ul of the Target Probe and incubated for 16 hours.

| Component | 10ng/20ng/30ng/50ng/100ng |
|---|---|
| TargetSeq One Hyb Buffe | 13ul |
| Hyb Human Block | 5ul |
| TargetSeq Eco Universal Blocking Oligo | 2ul |
| Rnase Block | 5ul |
| Total volume(add Nuclease-free Water) | 28ul |

**6.Post-capture nanopore library preparation**

In the post-capture nanopore library preparation step, cap beads are prepared and added to the hybridized product to selectively absorb the hybridized DNA fragments. After thorough washing, the cap beads with the absorbed DNA fragment are subjected to post-capture PCR under specific conditions.

| Component | 10ng-test1 | 20ng/30ng/50ng/100ng |
|---|---|---|
| previous products | 25ul | 25ul |
| Post PCR Master Mix | 2.5ul | 2.5ul |
| Post PCR Primer | 2.5ul | 2.5ul |
| PCR amplification processing | 95℃ 3min;<br>98℃ 20s;<br>60℃ 30s;<br>72℃ 1min; | 95℃ 1min;<br>98℃ 20s;<br>60℃ 30s;<br>72℃ 30s; |
| PCR cycle(different cycle for different input) | 10ng-1:18<br>10ng-2:20 | 16 |
| Processing | 72℃ 5min; | 72℃ 5min; |
| Purification(VAHTS DNA Clean Beads) | 1.1x | 1.1x |

After the PCR amplification step, the product is purified and end-repaird, then 300fmol product is used to construct the nanopore library following QLK-V1.1.1 protocol(QitanTech).

| Component | 10ng/20ng/30ng/50ng/100ng |
|---|---|
| cDNA | 300fmol |
| DRB | 7ul |
| EPM | 3ul |
| Total volume(add Nuclease-free Water) | 60ul |
| processing | 20℃10min;<br>65℃10min; |
| Purification(VAHTS DNA Clean Beads) | 1x |
| End-repaired products | 60ul |
| SAC | 5ul |
| LRB | 25ul |

| | |
|---|---|
| DLE | 10ul |
| hold still 10min | |
| Purification(VAHTS DNA Clean Beads) | 0.4x |
| AEB | 15ul |

**7.QNome nanopore sequence**
The 80fmol nanopore adapter ligated product is loaded into a nanopore cell for sequencing about 16h following the QSK-V1.1.1 protocol.

**8. Bioinfomatic analysis**
For illumina short read data, fastqc was uased to do quality control and star-fusion was used to detect fusions, the running example

```
singularity exec -e -B /path \
        /softpath/star-fusion.v1.12.0.simg \
        STAR-Fusion \
        --left_fq /datapath/left.fq \
        --right_fq /datapath/right.fq \
        --genome_lib_dir /softpath/source/ctat_genome_lib_build_dir \
        --no_remove_dups \
        -O /outpath/sample
```

For nanopore data, data quality control, reference alignment and LongGF fusion detection and filtering were described in our streamlined fusion detection pipeline(https://github.com/HuanYuu/TargetFusion). Running examples for other fusion detection method were described as follow.
Fusionseeker running example:

```
fusionseeker \
--bam /datapath/sample.sorted.bam \
--datatype nanopore \
--maxdistance 40 \
--minsupp 5 \
--thread 10 \
--keepfile \
--outpath /outpath/sample_fusionseeker \
--ref /referencepath/GRCh38.fa \
--gtf /referencepath/Homo_sapiens.GRCh38.104.chrname.gtf.gz
```

JAFFAL running example:

```
/softpath/tools/bin/bpipe run \
-n 10 \
/softpath/JAFFAL.groovy \
/fastqpath/sample.fq.gz
```

Genion running example:

```
genion \
-i sample.fq.gz \
--gtf /softpath/Genion_dependent/Homo_sapiens.GRCh38.97.gtf \
--gpaf /datapath/sample.sorted.paf \
-s /softpath/Genion_dependent/hg38_cdna.selfalign.tsv \
-d /softpath/Genion_dependent/genomicSuperDups.txt \
-o /outpath/sample.fusion.tsv \
--non-coding
```

Among the four softwares, LongGF got the highest recall rate(77.78%) and the result was more suitble for customized fitering(Supplementary Table 4).

Supplementary Table 4(sheet1): The fusion detection results for 7 libraries by 4 different softwares

| SampleID | TestID | Positive fusion pair | LongGF | fusionseeker | jaffal | genion |
|---|---|---|---|---|---|---|
| BCR-ABL1-QNome-short1 | YF0453 | BCR-ABL1 | BCR-ABL1 | BCR-ABL1 | BCR-ABL1 | - |
| BCR-ABL1-QNome-short1 | YF0454 | BCR-ABL1 | BCR-ABL1 | BCR-ABL1 | BCR-ABL1 | - |
| BCR-ABL1-QNome-long | YF1275 | BCR-ABL1 | BCR-ABL1 | - | BCR-ABL1 | - |
| CCPVS-SFQ | YF0711 | BCR-ABL1 | BCR-ABL1 | BCR-ABL1 | - | - |
| CCPVS-YZ | YF0710 | ETV6-PDGFRB | ETV6-PDGFRB | ETV6-PDGFRB | ETV6-PDGFRB | - |
| CCPVS-SFQ | YF0756 | BCR-ABL1 | BCR-ABL1 | - | BCR-ABL1 | - |
| CCPVS-YZ | YF0755 | ETV6-PDGFRB | ETV6-PDGFRB | - | ETV6-PDGFRB | - |
